# Impact of Seizure Detectors on Treatment and Costs of Epilepsy

**DOI:** 10.1101/2025.03.30.25324895

**Authors:** Michel J.A.M. van Putten, Maryam Amir Haeri

## Abstract

**Introduction:** Epilepsy management relies heavily on the accurate detection of seizures. Unreliable seizure readouts can lead to missed seizures and erroneous physician visits due to false positives. These inaccuracies can also result in delayed treatment or inappropriate medication adjustments, ultimately increasing healthcare costs. Here, we introduce a model to simulate the effects of an imperfect seizure readout on the (perceived) efficacy of treatment, the number of physician visits and associated health-care costs.

**Methods:** To assess the effects of the reliability of seizure detection systems on treatment, we introduce a simple model that generates seizures with a probability *P*, which can be altered by medication. In addition, we simulate different degrees of reliability of seizure detectors, where changes in medication are based on the *perceived seizure frequency*. We quantify the resulting costs in terms of remaining true seizures, physician visits, and the number of medication changes as a function of detector reliability.

**Results:** We show that the degree of unreliability in seizure readouts significantly affects overall healthcare costs. Costs increased up to three to five times more for unreliable systems. Moreover, unreliable systems may result in delayed or even inappropriate change of medication, further complicating effective treatment.

**Conclusion:** Reliable seizure detection systems can significantly reduce health-care costs and improve epilepsy treatment.

**Key Findings:**

- In a perfect detection system, costs arise primarily from true seizures, which include consultations with a neurologist and costs associated with hospital admissions or loss of productivity.
- As the unreliability of detectors increases, costs from missed seizures become more prominent, leading to untreated epilepsy episodes and limiting optimal patient care.
- Erroneous visits, resulting from unreliable seizure detection systems, may also contribute to suboptimal treatment and increase healthcare costs.

## 1 Introduction

Epilepsy is a neurological disorder characterized by an increased risk of recurring seizures, where treatment aims to reduce or prevent the occurrence of these events [3]. Reliable seizure detection is essential for evaluating the efficacy of medical interventions [3, 11]. Currently, seizure diaries are the primary method for tracking seizure frequency and evaluating treatment outcomes. While practical, these diaries have significant limitations, with both false negatives and false positives interfering with optimal care [4].

Underreporting is a major concern, often due to unrecognized seizures (e.g., those occurring during sleep or with impaired awareness). Additional factors include cognitive or motor impairments and reduced motivation to log events. Diaries may also misclassify non-epileptic events like psychogenic seizures, panic attacks, or syncopes. As a result, the sensitivity of seizure diaries can fall below 50%, especially for nocturnal seizures or those with impaired awareness [2, 7]. Even for tonic-clonic seizures, sensitivities remain suboptimal, with estimates around 50% [7].

Long term monitoring has revealed further discrepancies. In a study where ten patients recorded subcutaneous EEG (24/7 EEG™ SUBQ) for over 15 months [16], approximately 28% of diary-reported events were not actual seizures, indicating a specificity of 72%. Additionally, about 30% of the seizures detected by the device were not captured in diaries. Another study using this device, involving nine patients with epilepsy, found that agreement between the diary and the electrographic seizures was poor, with a mean Cohen’s kappa of 0.29, underscoring the limited reliability of seizure diaries [12].

While seizure diaries remain widely used, their inaccuracies have direct consequences on both patient care and healthcare expenditures. Missed seizures may delay treatment, increasing the risk of hospital admissions. Conversely, false positives can lead to unnecessary specialist visits and medication adjustments, further driving up costs [13].

Here, we quantify the financial impact of unreliable seizure readouts on the costs of epilepsy care using a simulation model. Our model assumes a baseline seizure likelihood, which can be modified by anti-seizure medication (ASM). The effectiveness of the medication is assessed based on the number of remaining seizures and the sensitivity and specificity of the seizure detection. Specifically, we quantify costs associated with missed seizures and false positive detections, considering their impact on hospital admissions, productivity loss, and unnecessary specialist visits. Our model reflects a realistic, real-world healthcare setting.

## 2 Methods

We introduce a simulation model to examine how inaccuracies in seizure readouts affect the costs of epilepsy care. The model is built around a baseline seizure likelihood which can be adjusted by anti-seizure medication (ASM). Additionally, it incorporates a mechanism for switching treatments when seizures persist despite medication, including due to false positive detections. A cost function is defined to capture the economic impact of both true and detected seizures, as well as physician visits and hospital admissions.

To explore how detection inaccuracies impact healthcare costs, we systematically vary the reliability of the seizure detector. This enables us to quantify how detection inaccuracies translate into increased healthcare expenses and suboptimal treatment outcomes. In our model, the likelihood of a seizure, *P*_seizure_ is defined by a prior probability based on the expected seizure frequency (e.g., 3 seizures per week). The impact treatment is captured by adjusting this likelihood according to

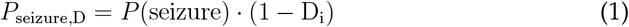

where *D*_*i*_ represents the effectiveness of the intervention on the seizure likelihood. *D*_*i*_ is in the range [0*−*1] where the subscript *i* refers to a particular drug. For instance, a perfect drug *D*_*A*_, that allows complete prevention of seizures, is characterised by *D*_*A*_ = 1, while another drug with no effect on the prevention of seizures is modelled as *D*_*B*_ = 0.

The occurrence of a seizure is determined stochastically by comparing this probability to a random value drawn from a uniform distribution in the interval [0 *−*1]. If the random value is smaller than the seizure probability a seizure occurs; otherwise, it does not. At each time step *t*, we thus draw a random value *r*_*t*_ from a uniform distribution and a seizure occurs if and only if

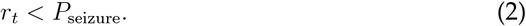

Effectiveness of treatment is estimated by the *perceived* reduction in seizure likelihood, estimated from the number of detected seizures (both true and false) occurring after treatment initiation. If, after the initial treatment, a seizure re-occurs (whether true or detected by the system), the neurologist will switch from drug *D*_*A*_ to *D*_*B*_. If the effect of *D*_*B*_ seems insufficient, treatment progresses to another switch, prescribing *D*_*C*_. These changes can also represent adjustments in drug dosage.

### 2.1 Cost Model

Seizures incur various costs, including hospital admissions, productivity losses, and physician consultations. False positive seizure detections often lead to unnecessary visits to a neurologist, while false negatives, i.e., missed or undetected seizures, also contribute to increased costs. These missed events can result in complications, reflecting suboptimal treatment and increasing the likelihood of future hospitalizations.

To model these costs, we define two components: costs associated with physician visits, *N*_visit_, and costs related to actual seizures, *N*_seizure_. The total cost of treatment *C*_total_ is a function of the number of physician visits due to both true *N*_visit,true_ and erroneous *N*_visit,false_ seizure detections, as well as the costs associated with the occurrence of true seizures *N*_seizure_, regardless of whether they are detected. This is expressed as

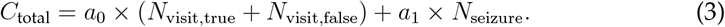

Here, *a*_0_ represents the cost of a single physician visit, and *a*_1_ represents the cost associated with each true seizure event, which we assume results in a consultation. We set = *a*_0_ = $340 and *a*_1_ = 5*a*_0_ = $1700, reflecting the assumption that the economic impact of a true seizure, including potential hospitalizations and productivity losses, is five times greater than the cost of a physician consultation. These numbers are derived from typical healthcare cost distributions in the U.S. as reported in a systematic review from 2014 [1]. It was estimated that an initial or follow-up consultation with a neurologist typically costs approximately $200, reflecting the median expense for specialist visits without additional diagnostics. A seizure requiring an ER visit was estimated at $2,000, covering tests and treatment. Productivity loss from missed work due to a seizure is associated with costs of approximately $250 per day, based on the average U.S. hourly wage, resulting in a total cost per seizure of $2,250, combining ER visit and productivity loss. Some seizures may also require an overnight stay in the hospital, adding even more costs. We adjusted for an average annual healthcare inflation rate of 5.5% and simplified and remained conservative, for which we set *a*_1_ to five times the costs of *a*_0_. This ensures that seizure-related costs are accounted for without overestimating their financial impact.

### 2.2 Modelling Reliability of the Seizure Detector

We introduce a seizure detector, with varying reliability, ranging from perfect (no false detections, sensitivity=100%) to maximally imperfect (random detections only).

We parametrise the reliability of our seizure detector by a variable *σ*, which represents the probability of misreporting. The probability of misreporting a seizure, a false positive (FP) is given by *σ*_1_, and the probability of failing to detect a seizure, a false negative (FN), is given by *σ*_2_. Our model allows us to simulate a range of possible detector reliabilities, where the sensitivity and specificity are given by

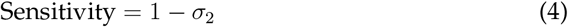

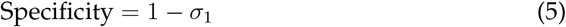

At each time step in the simulation, a seizure occurs with a probability determined by the effectiveness of the current drug treatment and the prior seizure likelihood. The probability of a seizure occurring is denoted as *P*_seizure_, which depends on the drug being used. If a seizure occurs, the detector’s reliability is modelled based on the value of *σ*, according to

- A perfect detector. Here, *σ*_1_ = *σ*_2_ = 0 any real seizure is always detected correctly, i.e., no false negatives occur.
- An unreliable detector. Here, *σ*_1_ *>* 0 or *σ*_2_ *>* 0, the probability of detecting a real seizure is 1 *− σ*_2_. If a seizure is missed, it is considered a false negative, increasing the number of missed seizures. If no seizure occurs, there is still a chance of a false positive to be generated with probability *σ*_1_.

### 2.3 Simulations

#### 2.3.1 Scenario I

In the first scenario, we assume a mean seizure frequency of *P*_seizure_ = 0.3 seizures per week. Further, we have three drugs available with efficacies of *D*_*A*_ = 0.5, *D*_*B*_ = 1, and *D*_*C*_ = 0.6. The physician starts treatment with drug *D*_*A*_, thus halving the likelihood of a seizure. A change in treatment will be made, switching to *D*_*B*_ if, after treatment initiation, a new seizure occurs. If the effectiveness of this drug is perceived as suboptimal, a final switch to the third drug occurs. The simulation flowchart is outlined in Fig. 1.

**Figure 1:**
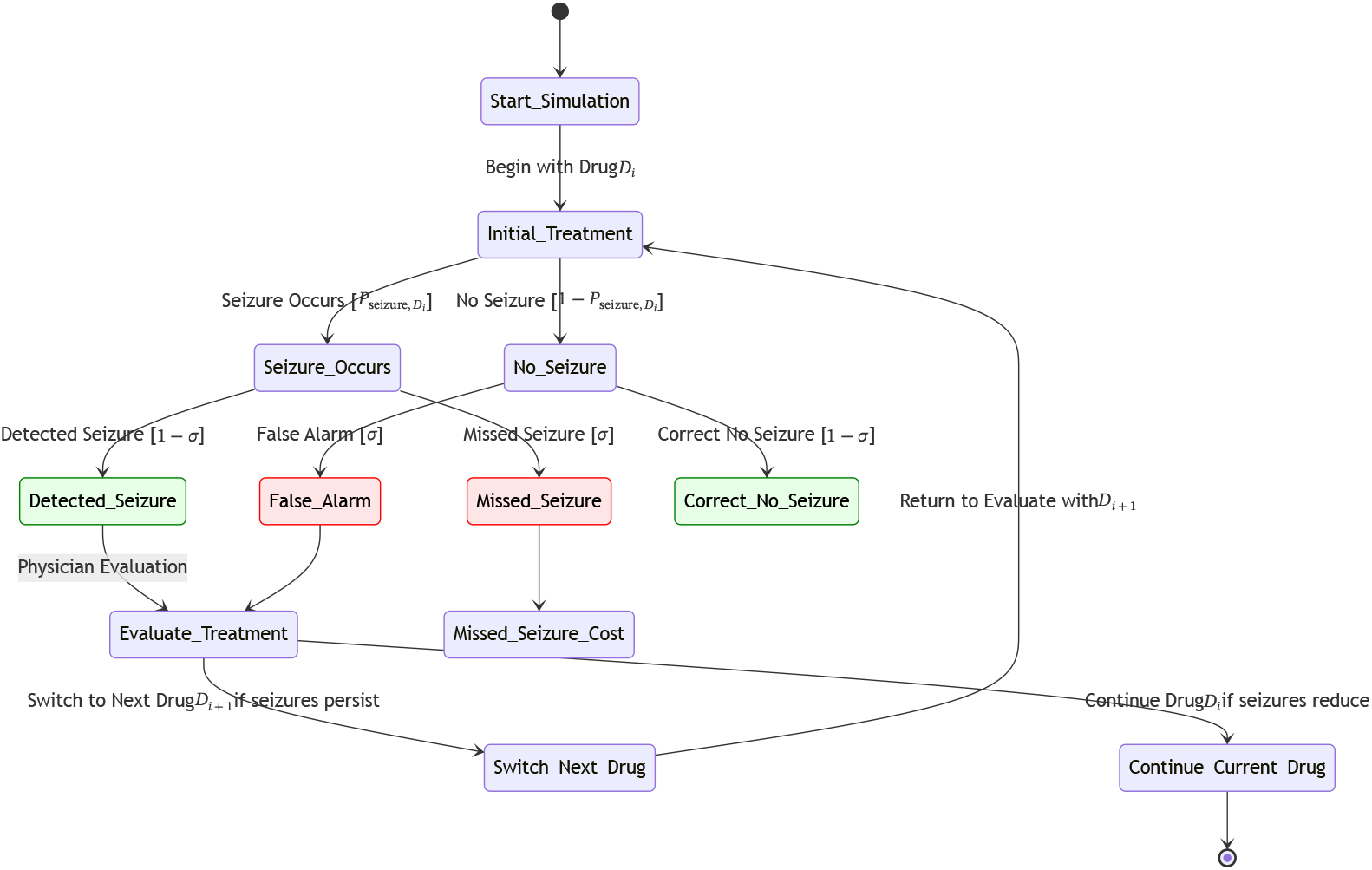
Flowchart of the simulation process for epilepsy treatment evaluation and seizure detection reliability. The simulation begins with an initial treatment using Drug *D*_*i*_. Depending on the probability of seizure occurrence (*P*_seizure,*D*_*i*), the system either detects or misses seizures. Green boxes represent correct detections or no-seizure outcomes, while red boxes indicate detection errors, including missed seizures and false alarms, influenced by the detector’s reliability (*σ*). Missed seizures contribute to untreated seizure costs, while false alarms lead to unnecessary physician visit costs.

The simulation was run for one thousand iterations with varying degrees of unreliability, according to four scenarios. First, we consider a perfect detector with negligible FPR (False Positive Rate) and FNR (False Negative Rate) (both *σ*_1_ = *σ*_2_ = 0). Second, we simulate a detector with equal imperfect sensitivity and specificity, with *σ*_1_ = *σ*_2_ in the range [0.05 *−* 0.3]. Next, we explore a detector characterized by low FPR, i.e. high specificity, but limited sensitivity (high FNR) with =*σ*_1_ = 0.05 and *σ*_2_ = 0.3). Finally, we simulate a detector with high sensitivity (low FNR) but poor specificity (high FPR) by setting *σ*_1_ = 0.3 and *σ*_2_ = 0.05).

By holding other simulation parameters constant and only varying these detection error rates, we can systematically assess how the different imbalances between sensitivity and specificity contribute to overall costs, physician visits, and missed seizure events. The cost components are calculated for each iteration and averaged across iterations. We also report on the average seizure frequency, the number of visits, and the number of drug switches. Details of the simulation are provided in Supplementary Material S1.

#### 2.3.2 Scenario II

Next, we simulate the clinical management of epilepsy over a two-year period (104 weeks) using a sequential drug-switching strategy with a generalized vector of drug effectiveness values. Treatment begins with the first drug and, whenever the cumulative count of real or perceived seizures exceeds a set threshold, the simulation switches to the next drug in the sequence. We employed a sequential drug-switching strategy using a drug effectiveness vector of {0.7, 0.7, 1.0, 0.7, 1.0, 0.7, 0.7, 0.7, 0.7, 0.7}, where the third and fifth drugs are assumed to be perfect (effectiveness = 1.0), while the remaining drugs reduce the baseline seizure probability by 30% (effectiveness = 0.7). Over a two-year period, the maximum number of drug switches or dosage modifications was set to nine.

Seizure occurrences are determined stochastically based on a baseline frequency, also set to 10 seizures per year, modified by the current drug’s effectiveness. Detection errors (false negatives and false positives) are incorporated via two detector settings: a perfect detector (*σ*_1_ = *σ*_2_ = 0.0) and an imperfect detector (*σ*_1_ = *σ*_2_ = 0.30). The simulation is run using 1000 Monte Carlo iterations. Weekly outcomes (physician visits, incurred costs, and drug switches) are cumulatively aggregated into monthly values to assess the overall impact of detection reliability on treatment outcomes and costs.

## 3 Results

With the perfect detector, after having started with the first drug *D*_*A*_ = 0.5 that reduces the likelihood of a seizure by 50%, a switch to the second ASM occurs due to incomplete seizure reduction. This will be correctly detected by the perfect detector, and treatment with a new drug, *D*_*B*_ = 1 is initiated, resulting in complete seizure reduction. In this scenario, the number of total seizures will be limited to n=2. Costs will be minimized, as the epilepsy is well-managed, and no false alarms will lead to unnecessary physician visits.

This contrasts with using an imperfect detector, with sensitivity = specificity less than 100%. While the medication change from *D*_*A*_ to *D*_*B*_ will also happen, the detector is imperfect. The erroneously detected seizures may induce a switch to drug *D*_*C*_, essentially resulting in a reduction of optimal treatment. This, however, goes unnoticed due to the imperfect detector. The net effect is more visits, both for erroneous detections and missed true seizures. Costs increase more than fivefold if sensitivities = specificities drop to around 70%. The metrics as a function of the detector unreliability are shown in Fig. 2.

**Figure 2:**
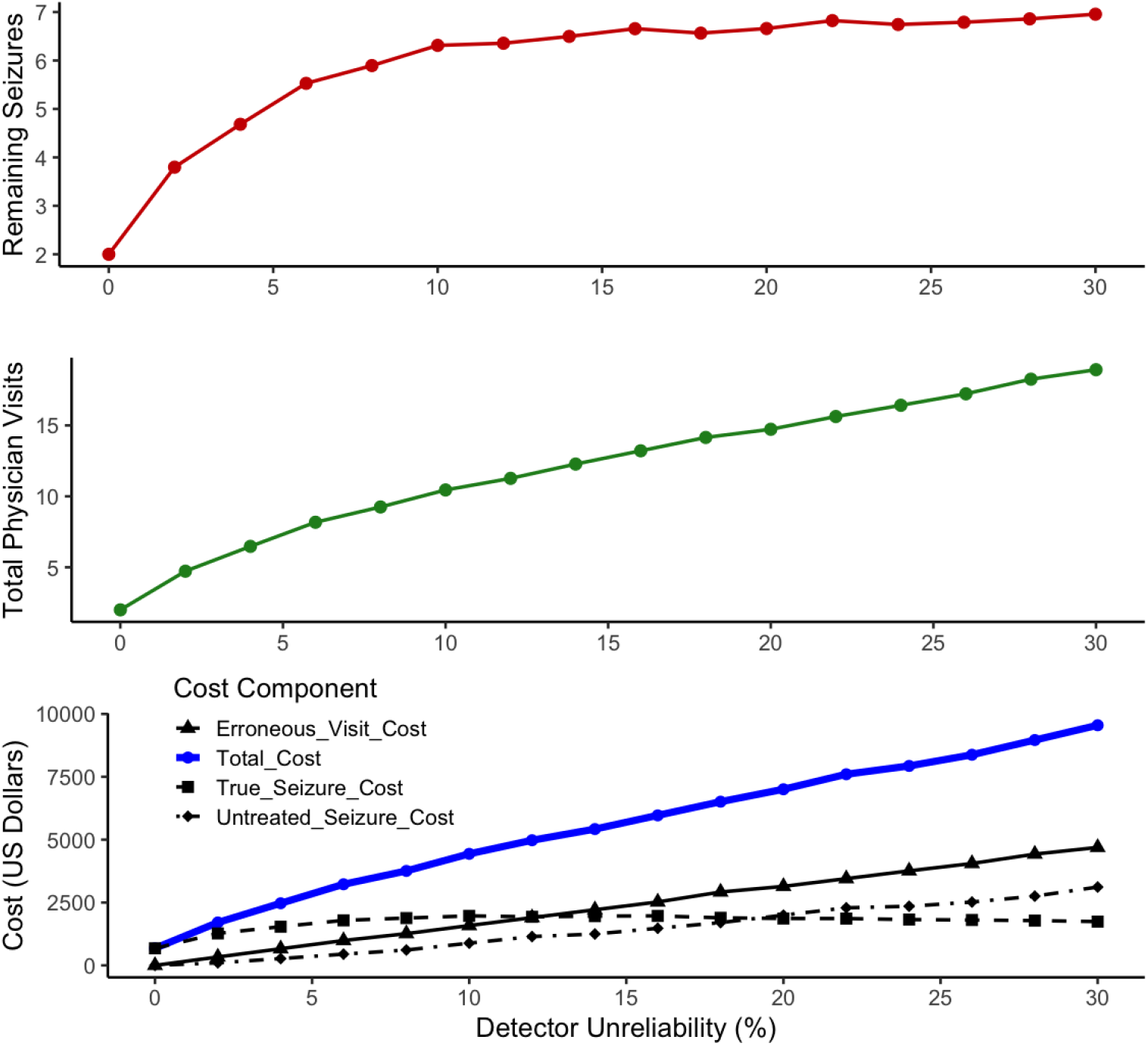
Number of seizures, visits and costs for increasing unreliability of the detector with equal sensitivity and specificity. Top: number of seizures as a function of the reliability of the detector. A reliability of 30% reflects a sensitivity=specificity of 70%. Middle: number of visits. Scenario A=0.5, B=1, C=0.6. Bottom: costs associated with visits, true and perceived seizures. The number of drug switches ranged from 1 (reliable system) to 2. Costs more than triple as the unreliability goes from 0 to 15% and increases even more than fivefold if *σ* = 0.3.

Next, we explored situations where the sensitivity and specificity of the detector differ, simulating either a good sensitivity but poor specificity or vice versa. The resulting outcome metrics are summarized in Table 1.

**Table 1:** Remaining Seizures and Total Physician Visits by Detector Reliability Profile. The number of seizures (both missed and correctly detected) increases with higher false negative rates, while the total physician visits indicate that a high false positive rate results in an increased number of clinical appointments due to erroneous seizure detections.

| Detector Reliability Profile | Number of Seizures | Total_Visits |
| --- | --- | --- |
| Perfect Detector | 2.00 | 2.00 |
| Low Sensitivity, High Specificity | 5.11 | 6.20 |
| High Sensitivity, Low Specificity | 6.82 | 20.36 |
| Low Sensitivity, Low Specificity | 7.03 | 18.97 |

As false negative rates increase, more real seizures go untreated, driving up the average number of remaining seizures. Conversely, scenarios with high false positives show that, while missed seizures may remain moderate, the overall detection inaccuracies still inflate the total number of seizures left unresolved. In addition, high false positives result in frequent, unnecessary visits, while high false negatives tend to keep visit counts lower but allow more actual seizures to go unaddressed.

Notably, the detector with “High False Positives, Low False Negatives” profile produces the greatest surge in total visits, as spurious detections keep prompting appointments. Together, these plots highlight how imbalanced detection errors—whether predominantly false positives or false negatives—significantly shape both clinical workload (physician visits) and patient outcomes (remaining seizures). An overview of the associated costs with various detector profiles is shown in Figure 3.

**Figure 3:**
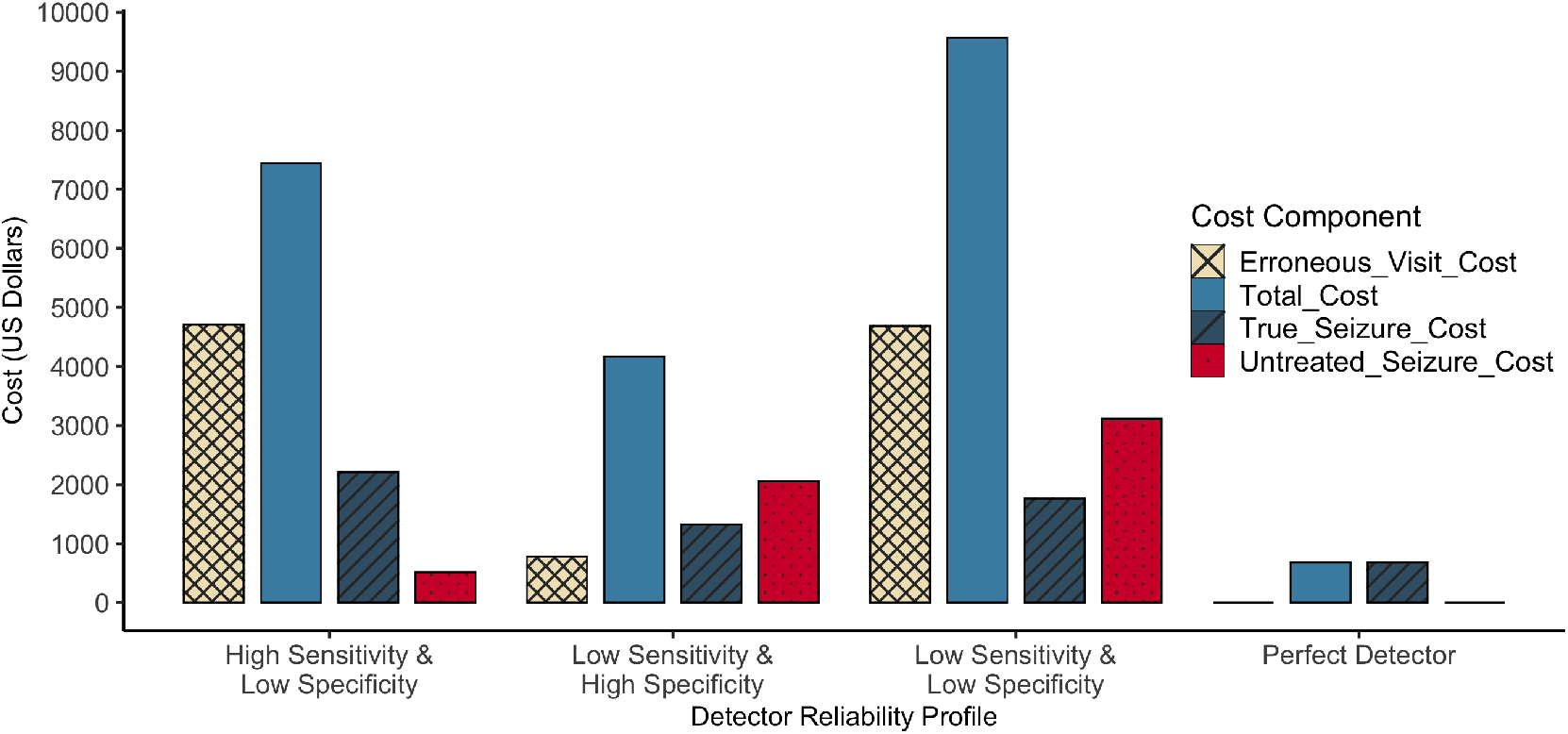
Average costs broken down by cost category for four detector profiles. Shown are costs for five scenarios with varying sensitivities and specificities, ranging from a perfect detector (left) to a detector with high FP and FN. Blue bars represent the total cost, gray bars indicate costs from true seizure visits, red bars show costs from untreated seizures, and orange bars show costs arising from erroneous visits.

**Figure 4:**
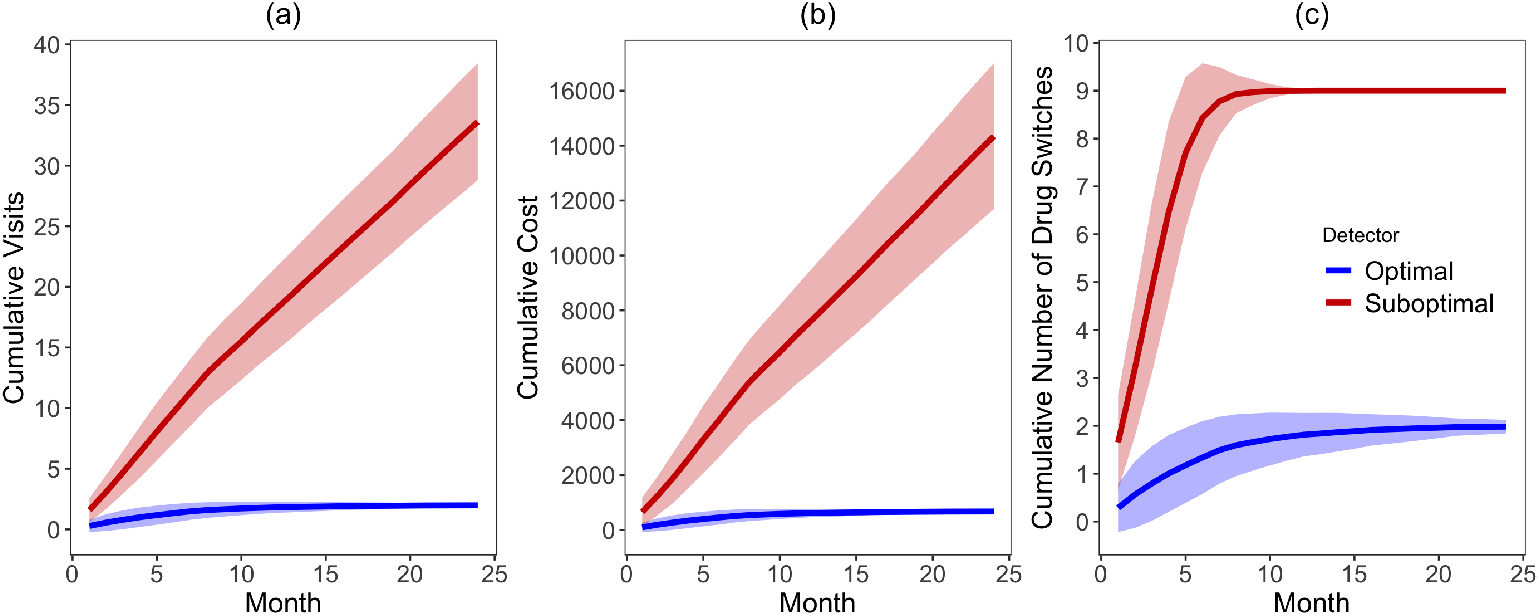
Cumulative monthly outcomes over two years for simulated epilepsy treatment using a sequential drug-switching strategy. The plots display cumulative physician visits, cumulative drug switches, and cumulative treatment costs, for two detector scenarios: optimal (*σ*= 0.0) and suboptimal (*σ* = 0.30). In this scenario, the third drug is assumed to be perfect, providing complete seizure suppression. Results are averaged over 1000 Monte Carlo runs.

Annual increase in costs for a detector with sensitivity and specificity of 70% (*σ* = 0.3) increases costs about five-fold. If we set annual costs in our model for a patient in a nearly perfect scenario to $2,000, assuming a single remaining seizure per year, we already observe a five-fold increase in costs due to the seizure detection system with sensitivity=specificity=85%, which is over $10,000.

## 4 Discussion

In this work, we address the effects of unreliable seizure detectors on healthcare costs. We show that as seizure detection becomes less reliable, costs from missed seizures and erroneous visits rise significantly. Assuming a sensitivity and specificity of 70%, which is likely optimistic compared to seizure diaries [7], the resulting costs can already be approximately five times higher than those associated with a perfect seizure detector. Based on our conservative cost estimates, this corresponds to approximately $4800 in additional expenses per year.

Our simulations show that even moderate levels of unreliability result in a substantial increase in missed seizures, reflecting suboptimal treatment and unnecessary physician visits. Additionally, a physician may erroneously decide to switch from ASM-B to a less effective ASM-C, resulting from a discrepancy between perceived and true seizure occurrences. In our example, although ASM-C is less effective, leading to an actual increase in the number of seizures, the unreliable readout fails to detect this. This underscores the critical need to improve the reliability of seizure readouts in epilepsy management.

Average annual costs for epilepsy patients, based on [1], inflation corrected, in the U.S. healthcare system are estimated to be approximately $13,000. Annual costs in our model for a patient in a nearly perfect scenario would be approximately only $2,000, where we assumed a remaining single seizure per year. Assuming the five-fold increase in costs due to unreliable seizure detection would amount to $10,000, which is of the same order of magnitude. We note that the costs for medication were not taken into account in our simulation that may range from $200 to over $2,000 per year.

While improving seizure readouts is crucial, the ideal scenario would involve proactive, real-time assessments of treatment efficacy, independent of seizures. For instance, measuring biomarkers, such as motivated by literature on excitatory/inhibitory (E/I) balance, could provide an indication of how well a treatment is working [5, 6, 14]. This could shift the focus from timely adjustments, enhancing patients’ outcomes. However, until such biomarker-driven approaches become clinically viable, reliable seizure detection systems remain a cornerstone of epilepsy management. Recent advancements include minimally invasive devices, such as subcutaneous implantable electrodes [17, 16], or in-ear electrodes, which offer long-term monitoring. For tonic-clonic seizures, commercially available options like the NightWatch [15] and Empatica Embrace [10] provide wearable solutions for home-based detection.

Our study has several limitations. First, we adopt a linear cost model with fixed expenses for physician contacts and seizures, even though actual costs may vary across regions and patient populations. Second, we assume a constant baseline seizure frequency, which does not reflect the circadian or multiday variations in seizure likelihood that have been documented in the literature [8, 9]. Third, we employ a threshold-based approach for drug switching, whereas in actual clinical practice, medication adjustments often involve additional considerations such as EEG findings, side-effect profiles, and patient-specific circumstances. Lastly, our simulations hold false positive and false negative rates at predefined values, while real-world detection systems may experience time-varying performance influenced by device adherence or environmental changes. Furthermore, one may reach the conclusion that the patient is pharmacoresistant, and that further treatment attempts to reduce the seizures are futile. Typically, however, at least two to three years pass before this conclusion is reached, also depending on the baseline seizure frequency.

Despite these limitations, our core message remains robust: reliability in seizure detection exerts a strong influence on healthcare costs and patient outcomes. Poor detection accuracy can drive up expenses by leading to both excessive, avoidable visits (from false positives) and serious complications resulting from unrecognized seizures (false negatives). Unreliable readouts, therefore, not only increase costs by leading to unnecessary physician visits and drug switches but may also result in suboptimal treatment changes that go unnoticed due to the system’s inability to accurately detect true events.

## Data availability

The code to generate the simulations will be made available on GitHub.

## S1: Simulation Procedure

### Simulation Steps

1. **Initialization**
  • Set the baseline seizure frequency: *P*_seizure_ = 0.3 seizures per week.
  • Initialize drug efficacy (*D*_*A*_ = 0.5, *D*_*B*_ = 1.0, *D*_*C*_ = 0.6).
  • Start treatment with drug *D*_*A*_.
  • Initialize counters:
    – *N*_seizure_ = 0 (missed seizures)
    – *N*_visit,true_ = 1 (initial physician visit)
    – *N*_visit,false_ = 0
    – *N*_drug switches_ = 0
    – Seizure count *C* = 0
2. **For each time step** *t* **(week) from 1 to 52:**
  (a) **Calculate the seizure probability under current drug:**

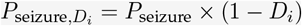
  (b) **Determine if a seizure occurs:**
    • Draw *r*_*t*_ *∼ U* (0, 1).
    • If *r*_*t*_ *< P*_seizure,*D*_*i*, a seizure occurs.
  (c) **Seizure detection:**
    • If a seizure occurs:
      – With probability 1 *− σ*, the seizure is detected (increment *N*_visit,true_ and *C*).
      – With probability *σ*, the seizure is missed (increment *N*_seizure_).
    • If no seizure occurs:
      – With probability *σ*, a false positive occurs (increment *N*_visit,false_ and *C*).
  (d) **Drug switching:**
    • If *C ≥ θ* (seizure threshold) and currently on *D*_*A*_:
      – Switch to drug *D*_*B*_.
      – Reset seizure count *C* = 0.
      – Increment *N*_drug switches_.
    • If *C ≥ θ* and currently on *D*_*B*_:
      – Switch to drug *D*_*C*_.
      – Reset seizure count *C* = 0.
      – Increment *N*_drug switches_.
3. **Cost calculation at the end of the simulation:** *C*_total_ = *a*_0_ *×* (*N*_visit,true_ + *N*_visit,false_) + *a*_1_ *× N*_seizure_

## Notes

### Competing Interest Statement

The authors have declared no competing interest.

### Funding Statement

This study did not receive any funding

## References

1. Charles E. Begley and Tracy L. Durgin. The direct cost of epilepsy in the United States: A systematic review of estimates. Epilepsia, 56(9):1376–1387, 2015.

2. Christian E Elger and Christian Hoppe. Diagnostic challenges in epilepsy: seizure under-reporting and seizure detection. The Lancet. Neurology, 17(3):279– 288, 2018.

3. Robert S. Fisher, Carlos Acevedo, Alexis Arzimanoglou, Alicia Bogacz, J. Helen Cross, Christian E. Elger, Jerome Engel, Lars Forsgren, Jacqueline A. French, Mike Glynn, Dale C. Hesdorffer, B. I. Lee, Gary W. Mathern, Solomon L. Moshé, Emilio Perucca, Ingrid E. Scheffer, Torbjörn Tomson, Masako Watanabe, and Samuel Wiebe. ILAE Official Report: A practical clinical definition of epilepsy. Epilepsia, 55(4):475–482, 2014.

4. Robert S. Fisher, David E. Blum, Bree DiVentura, Jennifer Vannest, John D. Hixson, Robert Moss, Susan T. Herman, Brandy E. Fureman, and Jacqueline A. French. Seizure diaries for clinical research and practice: limitations and future prospects. Epilepsy behavior : EB, 24(3):304–310, 2012.

5. Richard Gao, Erik J. Peterson, and Bradley Voytek. Inferring synaptic excitation/inhibition balance from field potentials. NeuroImage, 158(June):70–78, 2017.

6. Hai Yan He and Hollis T. Cline. What Is Excitation/Inhibition and How Is It Regulated? A Case of the Elephant and the Wisemen. Journal of Experimental Neuroscience, 13:10–12, 2019.

7. Christian Hoppe, Annkathrin Poepel, and Christian Elger. Epilepsy: Accuracy of Patient Seizure Counts. Archives of neurology, 64:1595–1599, 2007.

8. Philippa J. Karoly, Vikram R. Rao, Nicholas M. Gregg, Gregory A. Worrell, Christophe Bernard, Mark J. Cook, and Maxime O. Baud. Cycles in epilepsy. Nature Reviews Neurology, 17(5):267–284, 2021.

9. Marc G. Leguia, Ralph G. Andrzejak, Christian Rummel, Joline M. Fan, Emily A. Mirro, Thomas K. Tcheng, Vikram R. Rao, and Maxime O. Baud. Seizure Cycles in Focal Epilepsy. JAMA Neurology, 78(4):454–463, 2021.

10. Ming Zher Poh, Tobias Loddenkemper, Claus Reinsberger, Nicholas C. Swenson, Shubhi Goyal, Mangwe C. Sabtala, Joseph R. Madsen, and Rosalind W. Picard. Convulsive seizure detection using a wrist-worn electrodermal activity and accelerometry biosensor. Epilepsia, 53(5):93–97, 2012.

11. Julia C. M. Pottkämper, Jeannette Hofmeijer, Jeroen A. van Waarde, and Michel J.A.M. van Putten. The postictal state — What do we know? Epilepsia, 61(6):1045–1061, 2020.

12. Line S. Remvig, Jonas Duun-Henriksen, Franz Fürbass, Manfred Hartmann, Pedro F. Viana, Anne Mette Kappel Overby, Sigge Weisdorf, Mark P. Richardson, Sándor Beniczky, and Troels W. Kjaer. Detecting temporal lobe seizures in ultra long-term subcutaneous EEG using algorithm-based data reduction. Clinical Neurophysiology, 142:86–93, 2022.

13. Rafael Toledano, Vicente Villanueva, Manuel Toledo, Joel Sabaniego, and Paloma Pérez-Domper. Clinical and economic implications of epilepsy management across treatment lines in Spain: a real-life database analysis. Journal of Neurology, 270(12):5945–5957, 2023.

14. Anne M. van Nifterick, Danique Mulder, Denise J. Duineveld, Marina Diachenko, Philip Scheltens, Cornelis J. Stam, Ronald E. van Kesteren, Klaus Linkenkaer-Hansen, Arjan Hillebrand, and Alida A. Gouw. Resting-state oscillations reveal disturbed excitation–inhibition ratio in Alzheimer’s disease patients. Scientific Reports, 13(1):1–13, 2023.

15. Anouk van Westrhenen, Richard H.C. Lazeron, Johannes P. van Dijk, Frans S.S. Leijten, and Roland D. Thijs. Multimodal nocturnal seizure detection in children with epilepsy: A prospective, multicenter, long-term, in-home trial. Epilepsia, 64(8):2137–2152, 2023.

16. Pedro F Viana, Jonas Duun-henriksen, Andrea Biondi, J.S. Winston, D.R Freestone, A. Schulze-Bonhage, Benjamin H. Brinkmann, and Mark. P. Richardson. Real-world epilepsy monitoring with ultra long-term subcutaneous EEG: a 15month prospective study. medRxiv, (doi.org/10.1101/2024.11.16.24317163), 2024.

17. Pedro F. Viana, Jonas Duun-Henriksen, Martin Glasstëter, Matthias Dümpelmann, Ewan S. Nurse, Isabel P. Martins, Sonya B. Dumanis, Andreas Schulze-Bonhage, Dean R. Freestone, Benjamin H. Brinkmann, and Mark P. Richardson. 230 days of ultra long-term subcutaneous EEG: seizure cycle analysis and comparison to patient diary. Annals of Clinical and Translational Neurology, 8(1):288– 293, 2021.

